# Economic burden of Melanoma in Oregon to Inform State-wide Early Detection initiatives, The War on Melanoma

**DOI:** 10.1101/2025.09.26.25336709

**Authors:** Jeong-Yeon Cho, Jacob H. Nelson, Justin Ng, Tracy Petrie, Kyra Diehl, Elizabeth Stoos, Nathorn Chaiyakunapruk, Sancy Leachman

## Abstract

Melanoma imposes substantial clinical and financial burdens, particularly in advanced stages where systemic therapies are increasingly utilized. To establish a baseline for evaluating the statewide War on Melanoma™ (WoM) early detection initiative, we analyzed stage-specific melanoma expenditures in Oregon from 2011-2021 using a novel pseudostaging algorithm applied to the Oregon All Payer All Claims (APAC) database. The algorithm classified cases into early (in situ/localized), regional, and distant stages based on diagnostic, procedural, and treatment codes and was validated against SEER-Medicare claims and chart review. Over the study period, the number of melanoma cases in claims rose steadily, and total annual expenditures tripled, driven primarily by late-stage disease. Expenditures for early-stage melanoma remained relatively stable, while costs for regional and distant stages increased sharply with the adoption of immune checkpoint inhibitors and targeted therapies. These findings provide the first comprehensive, claims-based baseline of stage-specific melanoma costs in Oregon and highlight the disproportionate economic burden of advanced disease. Establishing such baselines is critical for evaluating population-level initiatives like WoM and for assessing the potential economic benefits of earlier detection and treatment.

**Summary:**

- We quantified the stage-specific economic burden of melanoma in Oregon using a pseudostaging algorithm applied to statewide claims data, demonstrating sharp increases in cost, particularly in advanced disease.
- These estimates provide essential baseline evidence for evaluating the War on Melanoma™, a statewide initiative designed to promote earlier detection and reduce the need for costly late-stage treatment.
- The reproducible claims-based approach offers a scalable framework for other states and health systems to assess the value of cancer prevention and early detection initiatives.

**Significance:** We establish stage-specific costs of melanoma in Oregon using a validated claims-based pseudostaging algorithm. The results highlight the disproportionate financial burden of advanced disease and provide essential context for the War on Melanoma™, a statewide early detection initiative. These baseline estimates enable rigorous evaluation of whether shifting toward earlier-stage diagnosis can reduce healthcare costs and improve patient outcomes, offering a framework relevant to other states and cancer prevention programs.

## Introduction

Melanoma is among the most lethal forms of skin cancer, with a rising incidence in the United States despite advances in prevention and treatment. In 2024 alone, more than 100,000 new melanoma cases were diagnosed nationally, with over 8,000 deaths attributed to the disease.^1^ Early-stage melanoma can often be treated effectively with simple surgical excision, leading to high survival rates and relatively low treatment costs. In contrast, although the advent of targeted and immunotherapies has improved patients’ survival in advanced-stage over the past decade, it requires more extensive and expensive treatments, which substantially increased healthcare expenditures.^2, 3^ The rapid expansion of novel melanoma therapies has dramatically reshaped both patient outcomes and healthcare costs. For example, among Medicare patients, the average first-year cost for stage IV melanoma increased by 61.7% between 2010 and 2015, largely due to immunotherapy adoption^4^. Total melanoma care costs among Medicare beneficiaries rose to more than $340 million in 2015, with most of this increase attributable to stage IV treatment. Moreover, additional approvals for earlier-stage adjuvant therapies further expand patient eligibility, amplifying long-term cost pressures. Beyond drug prices, the number of patients requiring treatment continues to grow. Adults treated annually for skin cancer rose from 4.9 million in 2007-2011 to 6.1 million in 2016-2018.^5, 6^

The War on Melanoma (WoM) is a statewide initiative in Oregon designed to reduce melanoma burden through education and early detection. By encouraging skin self-examinations (SSE) and enhancing provider diagnostic capacity, WoM aims to shift detection toward earlier, more treatable stages. Shifting detection from advanced stages to early stages has the potential to reduce morbidity, mortality, and treatment costs. Primary prevention and early detection have proven cost-saving in analogous contexts. The SunWise program is estimated to save $2-4 in medical care costs for every dollar invested.^7^ Similarly, detecting melanoma at earlier stages is far less expensive than treatment of advanced disease.^8^ The WoM was launched in Oregon since 2014, with major campaigns held in 2019 and 2022, with booster interventions in 2023 and 2024. Although there are several reports showing early promising results from WoM efforts on melanoma literacy and educational outcomes,^9–13^ its health economic impact is not yet evaluated due to the data lag time and on-going program efforts. Given the changing patterns of diagnostic and therapeutic options, expected survival, and healthcare expeditures, it is necessary to establish a clear baseline of melanoma burden and associated expenditures within a structured framework to assess whether statewide early detection program such as WoM can reduce costs while improving outcomes.

Moreover, the lack of comparable programs in neighboring states such as Washington and Utah creates a unique opportunity to evaluate the impact of Oregon’s War on Melanoma™ program as a natural experiment. These states share similar demographics, healthcare delivery systems, and access to National Cancer Institute (NCI)-designated cancer centers, yet Oregon uniquely launched a statewide early detection initiative in 2014.^11^ Establishing Oregon’s baseline burden of melanoma expenditures is therefore not only important for in-state policy but also lays the groundwork for future comparative analyses to assess whether large-scale public health interventions can alter the trajectory of melanoma costs and outcomes across states. This study aims to quantify trends in melanoma-related healthcare expenditures in Oregon from 2011 to 2021, stratified by disease stage, in order to establish a baseline and provide a foundation for evaluating whether statewide early detection efforts such as WoM can mitigate future financial and clinical burden.

## Methods

### Study setting and context

The War on Melanoma^TM^ integrates three primary components (details can be found in Supplementary Materials): public education campaigns to increase awareness and promote skin self-examinations (SSE), early detection training to enhance diagnostic accuracy, and improvements in surveillance through the linkage of clinical and claims data. Program infrastructure was developed between 2013 and 2019 through creation of registries, educational curricula for both the public and health professionals, and partnerships with public health agencies. These efforts were followed by a statewide media campaign beginning in 2019, emphasizing early detection and timely clinical follow-up. While this analysis does not evaluate the direct impact of WoM, the program provides important context for interpreting melanoma cost trends observed in Oregon.

### Oregon All Payer All Calims (APAC) database

The primary data source for the cost analysis was the Oregon All Payer All Claims (APAC) database, which has collected medical and pharmacy claims since 2011. APAC captures information for approximately 92% of Oregon residents across commercial insurers, and includes member demographics, diagnosis and procedure codes, prescription records, and amounts paid by both insurers and patients. This dataset is uniquely suited to estimating direct healthcare expenditures associated with melanoma at the population level. Melanoma-related claims were identified using International Classification of Diseases, Ninth and Tenth Revision (ICD-9/10) codes. Claims extraction encompassed diagnostic procedures such as skin biopsies and pathology reads, surgical treatments including wide excisions and Mohs procedures, sentinel lymph node biopsies, staging and follow-up imaging, radiation therapy, systemic therapies including chemotherapy, targeted therapy, and immunotherapy, as well as outpatient follow-up visits. Because basal and squamous cell carcinomas are not captured in cancer registries, we also identified related claims separately to provide context on biopsy and diagnostic activity.

### Identification of melanoma staging from claims

To complement claims-based cost estimates, melanoma incidence, mortality, and stage distribution were obtained from the Surveillance, Epidemiology, and End Results (SEER) Oregon registry. SEER data contain tumor-level variables such as stage at diagnosis, histology, and survival outcomes, and thus served as the reference for validating claims-based approaches to stage classification. Because APAC does not include disease staging information, we developed a claims-based pseudostaging algorithm to classify cases into “pseudo stages”. This algorithm was informed by melanoma treatment pathways and clinical practice. Early-stage, or “pseudostage 1&2” cases were defined by biopsy and excision without systemic therapy or lymph node dissection. Late-stage cases were identified by code indicating a patient received care such as systemic therapy, regional lymphadenectomy, radiation therapy, or surgery for distant metastases. Late-stage cases were then classified as “pseudostage 3” or “pseudostage 4” based on frequency of imaging. A full description of the algorithm and the code list is provided in Supplementary Appendix A.

### Analysis

The primary outcomes of the analysis were total and stage-stratified direct medical costs of melanoma, measured as the combined insurer and patient payments. We calculated costs per case by pseudostage on an annual basis and examined temporal changes in economic burden. Secondary outcomes included trends in diagnostic activity, particularly the frequency and costs of skin biopsies, pathology reads, and excisions. We analyzed claims data from 2011 to 2021, which covered the period preceding WoM initiation and the initial years of statewide activities. Costs were aggregated by calendar year and expressed in nominal dollars, with details of inflation adjustment provided in Appendix A. We stratified expenditures by pseudostage to assess cost differentials across disease severity. To examine temporal patterns, we conducted descriptive time-series analyses and calculated percent changes across three periods: pre-intervention baseline (2011–2013), early WoM program activities (2014–2018), and expanded public campaigns (2019–2021). This approach provides a comprehensive baseline assessment of melanoma expenditures and diagnostic intensity in Oregon, establishing the necessary platform for future evaluations of whether WoM achieves its intended goals of earlier detection, reduced late-stage diagnoses, and alleviation of cost burden.

## Results

### Overall trends in melanoma-related medical costs

Between 2015 and 2022, total medical costs for melanoma diagnosis and treatment in Oregon showed a consistent upward trajectory (Figure 1). Expenditures in 2015 were estimated to $15,992,162 USD, and while small year-to-year fluctuations were observed, a marked rise was observed during time frame. Therefore, the total nearly trippled over seven years, presenting $46,415,028 million USD by 2022. This overall pattern indicates a steady increase in the financial burden associated with melanoma across the decade, even after accounting for modest annual variability.

**Figure 1.**
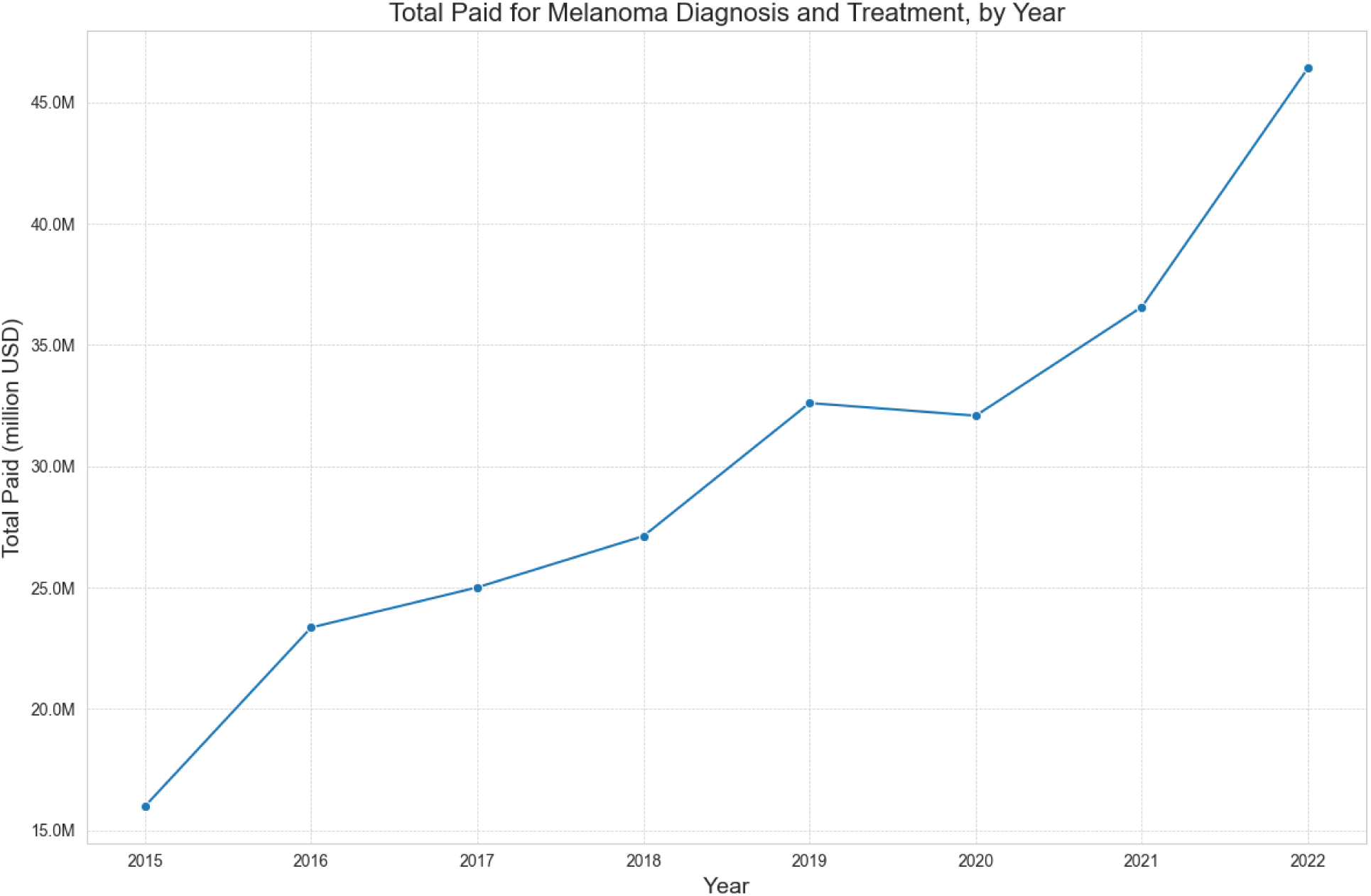
Total Costs for Melanoma Diagnosis and Treatment, by Year.

### Melanoma-related medical costs by disease stage

When disaggregated by melanoma stage defined by pseudostaging algorithm, expenditures for late-stage melanoma (defined as ‘pseudo 4’ by pseudostaging algorithm) were consistently greater than those for intermediate-stage (pseudo 3) and early-stage cases (Figure 2). In 2011, total medical costs at late-stage were estimated to 6,103,585 USD; by 2021, they had more than 4 times to about 25,168,420 USD. Total medical costs at intermediate-stage also increased substantially, rising from about 3,185,123 USD to nearly 8,307,895 USD during the same period. In contrast, early-stage melanoma expenditures remained relatively stable, ranging from 2,171,055 to 3,091,428 USD annually throughout the study window. By the end of the observation period, nearly half of all melanoma-related medical costs were attributable to late-stage disease. Analysis of average medical costs per patient further illustrates the substantial gradient in costs by melanoma stage (Figure 3). The average amount paid for a patient with late-stage melanoma was 124,719 USD compared with 19,893 for intermediate-stage and only $2,048 for early-stage disease. These averages highlight the disproportionate resource demands of advanced melanoma, with late-stage cases costing approximately 60 times more per patient than early-stage melanoma.

**Figure 2.**
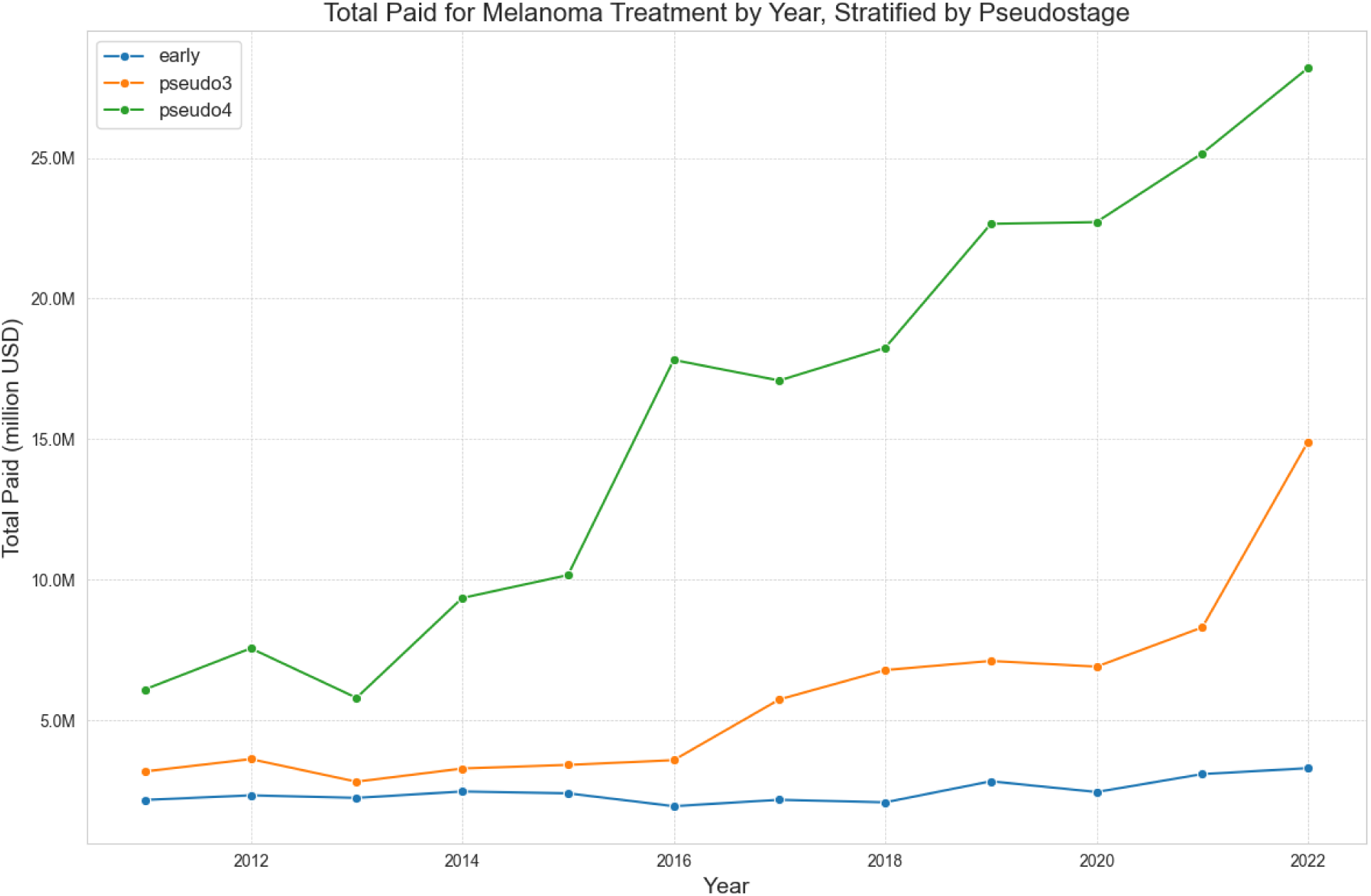
Cost of Melanoma Stratified by Pseudostage.

**Figure 3.**
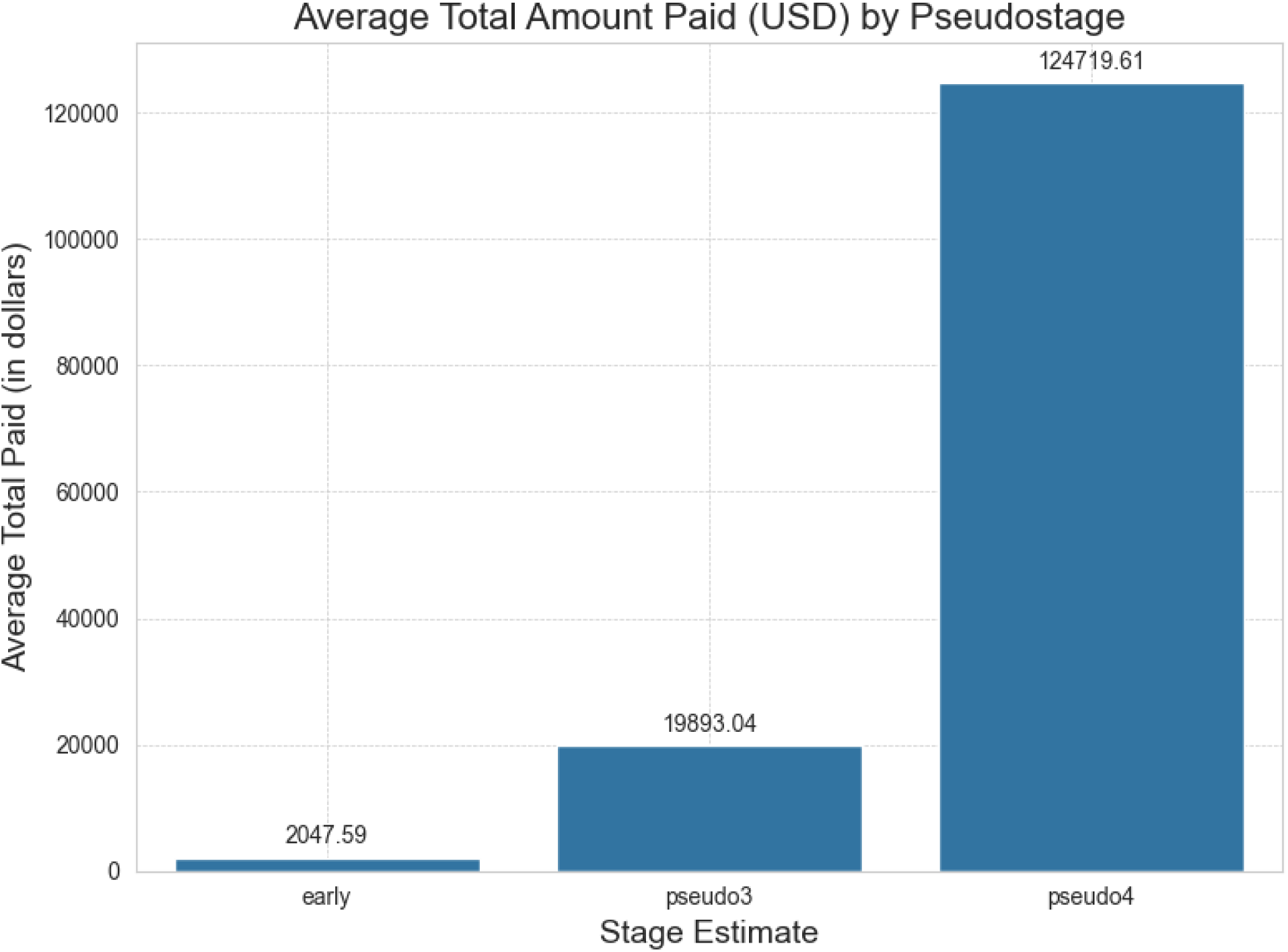
Average Cost of Melanoma Diagnosis and Treatment Stratified by Pseudostage.

### Biopsy utilization and costs

The volume of skin biopsy procedures remained high across the study period, averaging around 95,000 procedures annually (Figure 4). Procedure counts were relatively stable from 2011 through 2022. While short-term variability was evident, the overall trend suggests a persistent reliance on biopsy procedures as a diagnostic tool for melanoma in Oregon. Despite fluctuations in procedure counts, total payments for skin biopsy procedures remained remarkably stable, averaging approximately 8,494,911 USD annually between 2011 and 2021 (Figure 5). Modest increasing of total cost of biopsy was observed from 7,506,403 USD at 2011 to 10,526,723 USD at 2021. Taken together, Figures 4 and 5 suggest that while the costs of biopsies have increased, the average number of biopsies was relatively consistent across the decade.

**Figure 4.**
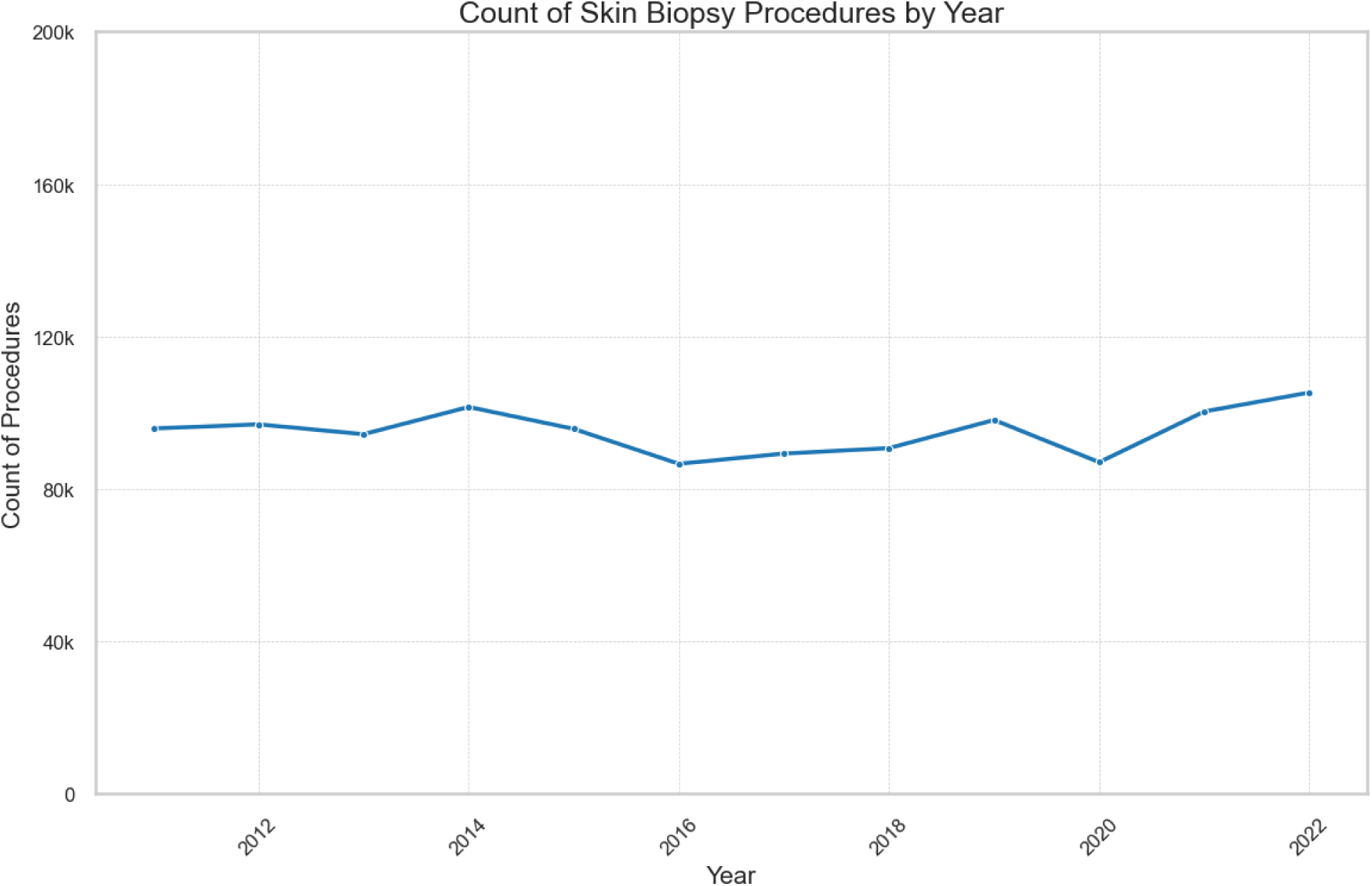
Count of Skin Biopsy Procedures by Year.

**Figure 5.**
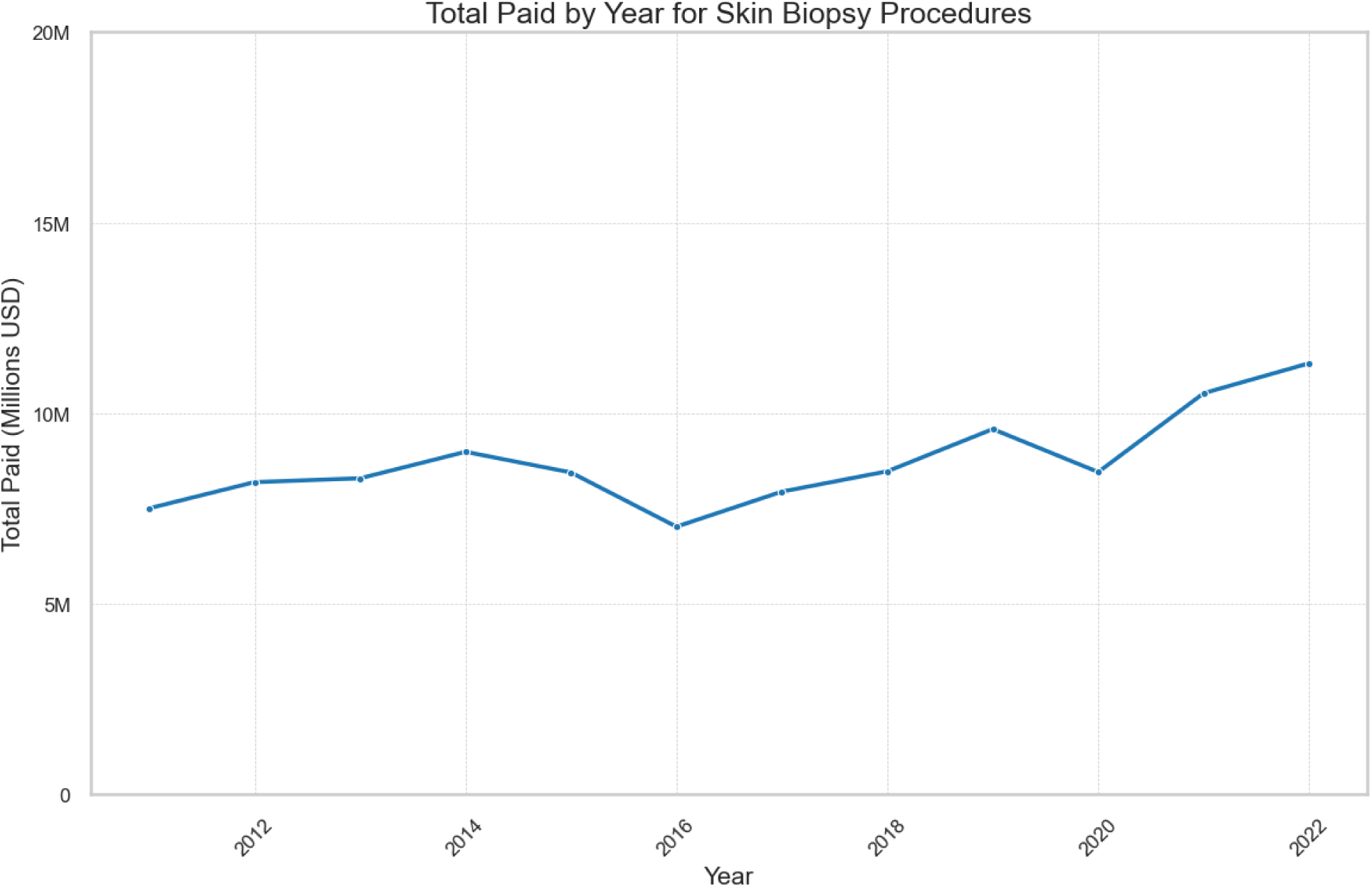
Total Paid for All Skin Biopsies, by Year.

## Discussion

Our analysis demonstrates a substantial rise in total payments for melanoma diagnosis and treatment in Oregon between 2015 and 2022, increasing from approximately 16 million USD to more than 46 million USD annually. This growth was most pronounced among patients with late-stage melanoma, where average payments exceeded 120,000 USD per case, compared to roughly 20,000 USD for intermediate-stage and just over $2,000 for early-stage melanoma. In contrast, the utilization of skin biopsy procedures remained relatively stable, averaging about 95,000 biopsies annually despite fluctuations. These findings highlight both the rising financial burden of melanoma care and the disproportionate costs associated with advanced disease.

These results underscore the importance of early detection. Identifying melanoma in localized stages enables curative surgical excision with minimal need for additional or advanced therapy, whereas advanced disease requires complex regimens including immunotherapy and targeted therapy, driving costs dramatically higher. Furthermore, the escalation of expenditures aligns with the approval and uptake of novel therapies, particularly immune checkpoint inhibitors and targeted agents. While these treatments have substantially improved survival outcomes, their high price and expanding indications from metastatic to stage III and now IIB/C disease have notably increased the overall financial burden of melanoma care.^2, 3^ Additionally, the stable costs of skin biopsies counter claims that melanoma is being overdiagnosed due to excessive diagnostic scrutiny.^14^ Recent concerns about the potential for overdiagnosis and increased diagnostic scrutiny have been raised due to SEER data revealing increased incidence of melanoma despite a relatively flat mortality rate.^14–17^ However, our real-world data from Oregon suggests that the number of biopsies performed statewide have been remarkably stable over similar periods of time, which does not support the hypothesis that increased rates of melanoma are due to increased diagnostic scrutiny. Concerns regarding unnecessary biopsies and overtreatment remain valid, but it is important to note that melanomas diagnosed at advanced stages, require costly systemic therapies that have significant side effects. Thus, future research should focus on linking biopsy rates, stage at diagnosis, and treatment expenditures to clarify these trade-offs.

Although our study provides population-level estimates that represents financial burden of melanoma in Oregon, several limitations should be acknowledged. Insurance claims data, including the Oregon All Payer All Claims (APAC) database, are subject to time lags of two to three years, coding limitations that obscure direct measurement of screening and skin examinations, and incomplete coverage due to the exclusion of Medicare fee-for-service and uninsured populations. Variability in payer submissions and the absence of linkage to cancer registry data further constrain precision. Additionally, the descriptive nature of this analysis limits causal inference, and the absence of formal comparison with neighboring states precludes evaluation of whether Oregon’s trends diverge from regional patterns. Another key limitation is the absence of a formal comparison with other states as a control group, which may limit the evaluation of whether Oregon’s trends diverge from regional patterns. Although our analysis include both pre- and post-periods of Oregon’s War on Melanoma™ initiative, comparative analysis with other states of similar size, demographics, and healthcare infrastructure (such as Washington and Utah, each anchored by a National Cancer Institute-designated cancer center) would provide valuable "natural experiments" to assess whether Oregon’s early detection emphasis translates into differences in cost trajectories and stage distribution. A baseline population survey comparing Oregon, Washington, and Utah demonstrated a similar demographic profile and baseline melanoma knowledge and behaviors across these three states,^11^ highlighting their suitability for future comparative health economic analyses.

## Conclusion

Our analysis highlights the rising economic burden of melanoma in Oregon, with costs driven predominantly by advanced-stage disease and the adoption of costly new therapies. Early detection remains the most promising strategy to mitigate both clinical and financial impact, offering the potential to reduce reliance on intensive late-stage interventions while improving patient outcomes. The War on Melanoma™ provides a model for leveraging public health initiatives to promote earlier detection, but further research-particularly comparative studies across states-will be necessary to confirm whether such efforts translate into measurable cost savings and improved survival at the population level.

## Data Availability

All data produced in the present work are contained in the manuscript. Patient-level data cannot be shared in accordance with Data Use Agreement.

## Supplementary Materials

## Appendix A. Program details for The War on Melanoma^TM^

The War on Melanoma (WoM) is a statewide initiative in Oregon aimed at reducing the burden of melanoma through earlier detection, improved education, and enhanced diagnostic capacity. Infrastructure development between 2013 and 2019 established the foundation of the program, including the creation of the Melanoma Community Registry with approximately 12,600 participants, the development of the MoleMapper mobile application which enrolled over 31,000 participants and generated more than 36,000 curated skin images, and the design of educational curricula for laypersons, primary care providers (PCPs), and skin service professionals.

Partnerships were also formed with the Oregon Health Authority, licensing boards, and other health organizations to support coordinated prevention and detection activities. During this period, WoM further expanded access through mobile screening events and teledermatology services, including E-visits and E-consults.

The program launched statewide media campaigns to increase public awareness and promote skin self-examinations. The initial 2019 “Start Seeing Melanoma” campaign was less effective and received mixed feedback, particularly in rural areas. In contrast, the 2022 “Melanoma Stands Out” campaign, developed by Oregon Health & Science University (OHSU), was better received and demonstrated improved cost-effectiveness by focusing on self-efficacy and clear early-detection messaging. A series of booster campaigns running from 2023 through 2027 refined messaging by targeting the media channels with the best return on investment.

WoM’s outreach activities were extensive and multi-sectoral. The program generated over 154 million media impressions across television, radio, and social media platforms and reached 13,000 high school students across 26 counties, resulting in measurable improvements in melanoma knowledge and skin check confidence. Engagement with the skin services industry produced more than 1,200 trained “Skin Crew” members and enrollment of 46 out of 150 beauty schools in the initiative. Among healthcare providers, WoM reached approximately 1,800 PCPs through Grand Rounds, distributed a PCP toolkit that was used by 829 clinicians, and improved diagnostic accuracy through structured training. Dermatology and melanoma specialists incorporated dermoscopy with teledermatology services, including E-visits and E-consults, achieving a 53% reduction in unnecessary in-person consultations.

## Appendix B Pseudostaging Algorithm

We developed a pseudostaging algorithm to classify melanoma cases into three categories-early (in situ/localized), regional, and distant-using claims data from the Oregon All Payer All Claims (APAC) database. Because tumor-level staging is not included in claims, the algorithm was informed by SEER data, clinical guidelines, and expert input. Validation steps included (1) manual review of ICD and CPT code dictionaries,, (2) chart review of 20 melanoma patients at Oregon Health & Science University to ensure coding consistency and face validity, and (3), validation with a supervised machine learning model.

Melanoma patients were identified using ICD-9/10 codes for malignant melanoma of the skin (C43.x) in primary or secondary diagnosis fields. Early stage (pseudo 1–2): Defined by biopsy and/or wide local excision without evidence of systemic therapy, lymph node dissection, or metastatic coding. Secondary diagnosis codes indicating metastasis (C77–C79) or palliative care (Z51.5) were also incorporated to capture advanced disease. This algorithm produced three consolidated pseudostage categories: Pseudostage 1&2 (Early/Localized): Biopsy and excision only; Pseudostage 3 (Regional): Evidence of regional spread, lymphadenectomy and/or systemic therapy. Pseudostage 4 (Distant): Evidence of metastasis, systemic therapy, and intensive imaging schedule (imaging events at 90 day intervals +/- 10 days).

The deterministic classification above demonstrated high concordance with a supervised machine learning model applied to the same dataset, and cases confirmed the ability of the pseudostaging algorithm to approximate SEER Summary stages using claims data.

**Supplementary Table B1.** Pseudostaging criteria.

| **Pseudostage** | **Criteria** | **Key Codes** | **Example Case** |
| --- | --- | --- | --- |
| **MIS, Stage I/II**<br>**(Localized)** | Wide local excision (WLE) or sentinel lymph node biopsy (SLNB) present **and** no systemic therapy | CPT: 116xx (WLE) | Excision of malignant lesion, no systemic therapy → Stage I/II |
| **Stage III**<br>**(Regional)** | Wide local excision **and** lymph node dissection or systemic therapy | CPT: 116xx, 387xx (lymph node dissection) | Excision + lymphadenectomy → Stage III |
| **Stage IV**<br>**(Distant/metastatic)** | ICD/CPT evidence of metastatic disease **or** systemic therapy (chemo/immunotherapy) | ICD-10: C77–C79 (metastatic sites); CPT: Drugs: J9xxxx (e.g., ipilimumab J9228), imaging codes (7xxxx) | Excision + systemic therapy (e.g., ipilimumab) + PET imaging every 3 months → Stage IV |

**Supplementary Table B2.** Diagnosis codes (ICD-10) used to identify melanoma and stage-related conditions.

| **Code(s)** | **Description** | **Used For** |
| --- | --- | --- |
| C43.0-C43.9 | Malignant melanoma of skin (by body site) | Case identification |
| C77.0-C77.9 | Secondary and unspecified malignant neoplasm of lymph nodes | Regional stage |
| C78, C79 | Secondary malignant neoplasms of lung, brain, bone, other sites | Distant stage |
| Z85.82 | Personal history of malignant melanoma | Exclusion/validation |

**Supplementary Table B3.** Procedural codes (CPT/HCPCS) for melanoma-related care.

| **Code(s)** | **Description** | **Used For** |
| --- | --- | --- |
| 11102–11107 | Tangential, punch, incisional/excisional biopsies | Biopsy flag |
| 11600–11646 | Excision, malignant lesion (by size/location) | Local excision |
| 38720, 38500 | Lymph node excision/dissection | Regional disease proxy |
| 36591, 36592 | Central venous catheter placement for infusion | Advanced therapy proxy |

**Supplementary Table B4.** Systemic therapy (NDC/RxNorm) proxies for distant stage indicator.

| **Drug Class** | **RxNorm Codes** | **Brand/Generic** |
| --- | --- | --- |
| Immune checkpoint inhibitors | 1792771 | Pembrolizumab (Keytruda) |
|  | 1792773 | Nivolumab (Opdivo) |
| BRAF/MEK inhibitors | 1373464 | Dabrafenib |
|  | 1796956 | Trametinib |
| CTLA-4 inhibitor | 1792775 | Ipilimumab (Yervoy) |

## Notes

### Competing Interest Statement

The authors have declared no competing interest.

### Funding Statement

This project was supported by the Cancer Early Detection Advanced Research Center (CEDAR) (CEDAR Project ID# Full 2022-1525) at OHSU, OHSU Knight Cancer Institute NCI Cancer Center Support Grant 2P30CA069533-24 and generous donations made possible through the OHSU.

### Author Declarations

This study used secondary de-identified claims databases from Oregon Health Authority.

